# Genome-wide analyses reveal novel opioid use disorder loci and genetic overlap with schizophrenia, bipolar disorder, and major depression

**DOI:** 10.1101/2022.09.09.22279755

**Authors:** Børge Holen, Alexey A. Shadrin, Romain Icick, Guy Hindley, Linn Rødevand, Kevin S. O’Connell, Oleksandr Frei, Shahram Bahrami, Weiqiu Cheng, Nadine Parker, Markos Tesfaye, Piotr Jahołkowski, Naz Karadag, Anders M. Dale, Srdjan Djurovic, Olav B. Smeland, Ole A. Andreassen

## Abstract

Opioid use disorder (OUD) and mental disorders are often comorbid, with increased morbidity and mortality. The causes underlying this relationship are poorly understood. Although these conditions are highly heritable, their shared genetic vulnerabilities remain unaccounted for. We applied the conditional/conjunctional false discovery rate (cond/conjFDR) approach to analyse summary statistics from independent genome wide association studies of OUD, SCZ, BD and MD. Next, we characterized the identified shared loci using biological annotation resources. OUD data was obtained from the Million Veteran Program (15,756 cases 99,039 controls). SCZ (53,386 cases 77,258 controls), BD (41,917 cases 371,549 controls) and MD (170,756 cases 329,443 controls) data was provided by the Psychiatric Genomics Consortium. We discovered genetic enrichment for OUD conditional on associations with SCZ, BD, MD and vice versa, indicating polygenic overlap with identification of 14 novel OUD loci at condFDR<0.05 and 7 unique loci shared between OUD and SCZ (n=2), BD (n=2) and MD (n=7) at conjFDR<0.05 with concordant effect directions, in line with estimated positive genetic correlations. Two loci were novel for OUD, one for BD and one for MD. Three OUD risk loci were shared with more than one psychiatric disorder, at *DRD2* on chromosome 11 (BD and MD), at *FURIN* on chromosome 15 (SCZ, BD and MD), and at the major histocompatibility complex region (SCZ and MD). Our findings provide new insights into the shared genetic architecture between OUD and SCZ, BD, and MD, indicating a complex genetic relationship, suggesting overlapping neurobiological pathways.

## 1 INTRODUCTION

Opioid use disorder (OUD) causes substantial morbidity and mortality worldwide ^1^. Some countries are more affected, such as the USA with the opioid crisis ^2^. Although OUD is less prevalent than other substance use disorders, OUD is an enormous public health burden with high rates of overdose deaths, which increased during the COVID-19 pandemic ^3^.

OUD is relatively prevalent in patients with severe mental disorders, such as schizophrenia (SCZ), bipolar disorder (BD) and major depression (MD) ^4-6^, all of which are associated with affective and psychotic symptoms to various degrees. Comorbidities lead to greater suffering compared to having either disorder alone ^7^, and often impede treatment ^4^. In patients with severe mental disorders the estimated prevalence of co-occurring OUD was 2.6-5.3% ^4^. Among adults with OUD the prevalence of a co-occurring severe mental disorder was estimated to be 27 % ^8^. At a sub-diagnostic threshold, an increased risk of nonmedical opioid use was reported for patients who had already been diagnosed with a severe mental disorder ^9^. Similarly, an increased risk of developing severe mental disorders for patients with existing nonmedical opioid use was also found ^9^. Further, there are several lines of evidence suggesting an overlapping neurobiological substrate related to the reward system and dopamine across mental illness in general ^10^, OUD ^11^, mood disorders ^12^, and psychotic disorders ^13^. E.g., can negative symptoms in SCZ, a category of internal heterogeneity, be conceptualized as a reward processing impairment ^14^. Better understanding of the mechanisms underlying these comorbidities is crucial for improving treatment and quality of life of affected individuals.

OUD and severe mental disorders can increase the risk of developing one another, have common environmental and genetic risk factors ^15^. Both OUD and severe mental disorders have moderate to high heritability estimates from twin and family studies, with OUD at 0.50 ^16^ SCZ at 0.80 ^17^, BD at 0.70 ^18^ and MDD at 0.35 ^19^. Recent progress in genotyping technology and international collaborations assembling large genome-wide association studies (GWAS) have provided novel insights into the genetic architecture of these complex disorders. A key discovery is that these disorders are highly polygenic ^20^, i.e., associated with many genetic risk variants, each with a small effect. Genetic studies have also suggested shared genetic aetiologies across severe mental disorders ^15^, including recent research which has found significant concordant genetic correlations (r_g_) between substance use disorders and severe mental disorders ^16^ with genetic correlation estimates between OUD and SCZ at 0.29, for BD at 0.16 and MD at 0.35 ^21^, suggesting common genetic risk factors. However, genetic correlation is a genome-wide measure, which includes the effects of all Single Nucleotide Polymorphisms (SNPs), and thus does not identify overlap at the individual locus level. Moreover, genetic correlation is unable to capture genetic overlap in the presence of shared genetic variants with a mixture of same and opposite effect directions as they “cancel each other out” ^22^. In recent years, genome-wide analyses demonstrated genetic overlap with mixed effect directions among a wide range of human traits and disorders, regardless of their genetic correlations ^23^. Therefore, investigating genetic overlap beyond genetic correlation is required to further elucidate the genetic architecture of complex disorders and their genetic relationships. Here, we aimed to reveal more of the shared genetic architecture between OUD and the mental disorders SCZ, BD and MD by applying analytical tools designed for polygenic architectures ^24^. We applied the conditional/conjunctional false discovery rate (cond/conjFDR) tools to boost discovery of genetic variants and to identify overlapping genetic loci between OUD, SCZ, BD and MD, beyond genetic correlation ^24^.

## 2 MATERIALS AND METHODS

### 2.1 GWAS Samples

Independent GWAS data were obtained in the form of summary statistics (p-values and effect sizes) (Table 1.). For OUD, data from the Million Veteran Program (MVP), Yale-Penn and the Study of Addiction: Genetics and Environment (SAGE) ^21^ were used. In the MVP cohort, case and control status were defined using electronic health record–data. There were 8,529 affected European American (EA) individuals and 71,200 opioid-exposed EA controls, 4,032 affected African American (AA) individuals and 26,029 opioid-exposed AA controls. The Yale-Penn and SAGE datasets included 2,015 EA cases and 963 controls and 1,180 AA cases and 847 controls from a previous GWAS ^25^. To ensure compatibility of linkage disequilibrium (LD) pattern, our main analysis on OUD focused on individuals of EA ancestry and included a total number of 10,544 cases and 72,163 opioid-exposed controls. GWAS data on SCZ (53,386 cases and 77,258 controls) ^26^, BD (41,917 cases and 371,549 controls) ^27^ and MD (170,756 cases and 329,443 controls) ^28^ were obtained from the Psychiatric Genetics Consortium (PGC) and were of European ancestry. See Supplementary methods for details.

**Table 1.** Genome-wide association studies used in the present analyses. OUD = opioid use disorder; SCZ = schizophrenia; BD = bipolar disorder; MD = major depression; MVP = Million Veteran Program; PGC = Psychiatric Genomics Consortium.

| Disorder | Analysis | Consortium | Sample size, n | Ancestry | SNPs, n | Reference |
| --- | --- | --- | --- | --- | --- | --- |
| OOD | Discovery | MVP | 10,544 cases<br>72,163 controls | European | 5,070,000 | 21 |
| SCZ | Discovery | PGC | 53,386 cases<br>77,258 controls | European | 7,585,078 | 40 |
| BD | Discovery | PGC | 41,917 cases<br>371,549 controls | European | 7,608,183 | 27 |
| MD | Discovery | PGC | 170,756 cases<br>329,443 controls | European | 7,666,894 | 28 |
| OOD | Validation | MVP | 5,212 cases<br>26,876 controls | African<br>American | 6,910,000 | 21 |
| SCZ | Validation | PGC | 22,778 cases<br>35,362 controls | East Asian | 9,607,215 | 32 |
| BD | Validation | FinnGen | 4,501 cases<br>192,220 controls | European | 14,114,100 | Publicly available<br>(https://r5.finnngen.fi/) |
| MD | Validation | 23andMe | 75,607 cases<br>231,747 controls | European | 13,519,496 | 33 |

### 2.2 Statistical Analyses

We visualized cross-trait enrichment using conditional quantile-quantile (Q-Q) plots, where distribution of p-values of all SNPs of a primary phenotype and for strata defined by the p-values for association with a secondary phenotype are represented. Cross-trait enrichment is evident if enrichment of statistical associations in the primary phenotype increases with increased significance of association with the secondary phenotype ^29^. We then applied the conditional FDR (condFDR), which leverages cross-trait enrichment between two phenotypes to improve genetic discovery. CondFDR readjusts the test-statistics in the primary phenotype by conditioning on SNP associations with the secondary phenotype, returning a condFDR value. OUD loci were identified at condFDR<0.05. We then applied the conjunctional FDR (conjFDR) tool to increase discovery of shared genetic loci jointly associated with OUD and in turn SCZ, BD and MD. The conjFDR approach is an extension of condFDR ^29^. Reversing the order of the phenotypes gives the condFDR value for the second phenotype conditioned on the first phenotype. ConjFDR then identifies SNPs which are significantly associated with both phenotypes ^24^. Shared loci were found at conjFDR<0.05, in line with prior literature ^30^. To control for spurious enrichment, random pruning was averaged over 500 iterations, and one SNP in each LD block (r^2^>0.1) was randomly selected for each iteration. We excluded SNPs within the major histocompatibility complex (MHC) (chr6:25000000-33000000), the chromosomal region 8p23.1 (location 7200000-12500000) and the gene *MAPT* (chr17:40000000-47000000) before fitting the FDR model given their complex LD structures that may bias FDR estimation ^31^.

We performed conjFDR analyses in independent samples for OUD (5,212 cases and 26,876 controls) ^21^, SCZ (22,778 cases and 35,362 controls) ^32^, BD (4,501 cases and 192,220 controls) [https://r5.finngen.fi/], and MD (75,607 cases and 231,747 controls) ^33^ (Table 1). See Supplementary methods for details. We then conducted a version of the binomial test, i.e., sign concordance test ^34^. Finally, we tested for variants nominally significant at p<0.05 in each independent sample. All analyses were conducted in Oslo, Norway.

### 2.3 Genetic locus definition

We defined independent genetic loci according to the FUMA protocol ^35^. Briefly, independent significant genetic variants were identified as variants with conjFDR<0.05 and LD r2<0.6 with each other. A subset of these independent significant variants with LD r2<0.1 were then selected as lead variants. For each lead variant all candidate variants were identified as variants with LD r2≥0.6 with the lead variant. For a given lead variant the borders of the genetic locus were defined as min/max positional coordinates over all corresponding candidate variants. Loci were then merged if they were separated by less than 250kb. LD information was calculated from the 1000 Genomes Project European-ancestry reference panel ^36^. Directional effects of loci were evaluated by comparing z-scores of the lead SNPs. We investigated all discovered loci for overlap with previously identified loci using our internal database which includes updated GWAS on all the traits investigated in this study.

### 2.4 Functional annotation of loci shared between OUD and severe mental disorders

The FUMA application SNP2GENE was run to identify genes mapped to the candidate SNPs with a conjFDR value <0.1 ^35^. All analyses were corrected for multiple comparisons. To minimize false positive and negative findings, we implemented a gene mapping strategy of qualification of at least two out of three of the following criteria: physical proximity of SNPs to genes, expression Quantitative Trait Loci (eQTL) and chromatin interaction mapping. SNP deleteriousness was measured by a Combined Annotation-Dependent Depletion (CADD)-score above 12.37 ^37^. For complementarity we also used the open-source Open Targets Genetics application, Variant to Gene (V2G) ^38^ to map lead SNPs to genes. V2G utilizes physical proximity of SNPs to genes, molecular phenotype quantitative trait loci investigations (QTL) which maps SNPs to genes where expression level is influenced by allelic variation at the SNP level (eQTL), and protein Quantitative Trait Loci (pQTL) which maps SNPs to genes where protein level is influenced by allelic variation at the SNP level, and chromatin interaction where 3D DNA-DNA interactions are considered. V2G leverages this information in machine learning algorithms on the input of lead SNPs.

## 3 RESULTS

### 3.1 Cross-trait enrichment

To visualize cross-trait enrichment, we constructed stratified quantile-quantile (Q-Q) plots that show successive increments of SNP enrichment for OUD conditional on increasing levels of SNP associations with SCZ, BD and MD (Figure 1 A-C). We then reversed the stratified Q-Q plots displaying the SNP associations for SCZ, BD and MD conditional on SNP associations with OUD (Figure 1 D-F). In all cases we observed successive upward and leftward deflection of the Q-Q plots after conditioning the primary phenotype on the conditional phenotype, indicative of genetic overlap between the phenotypes.

**Figure 1.**
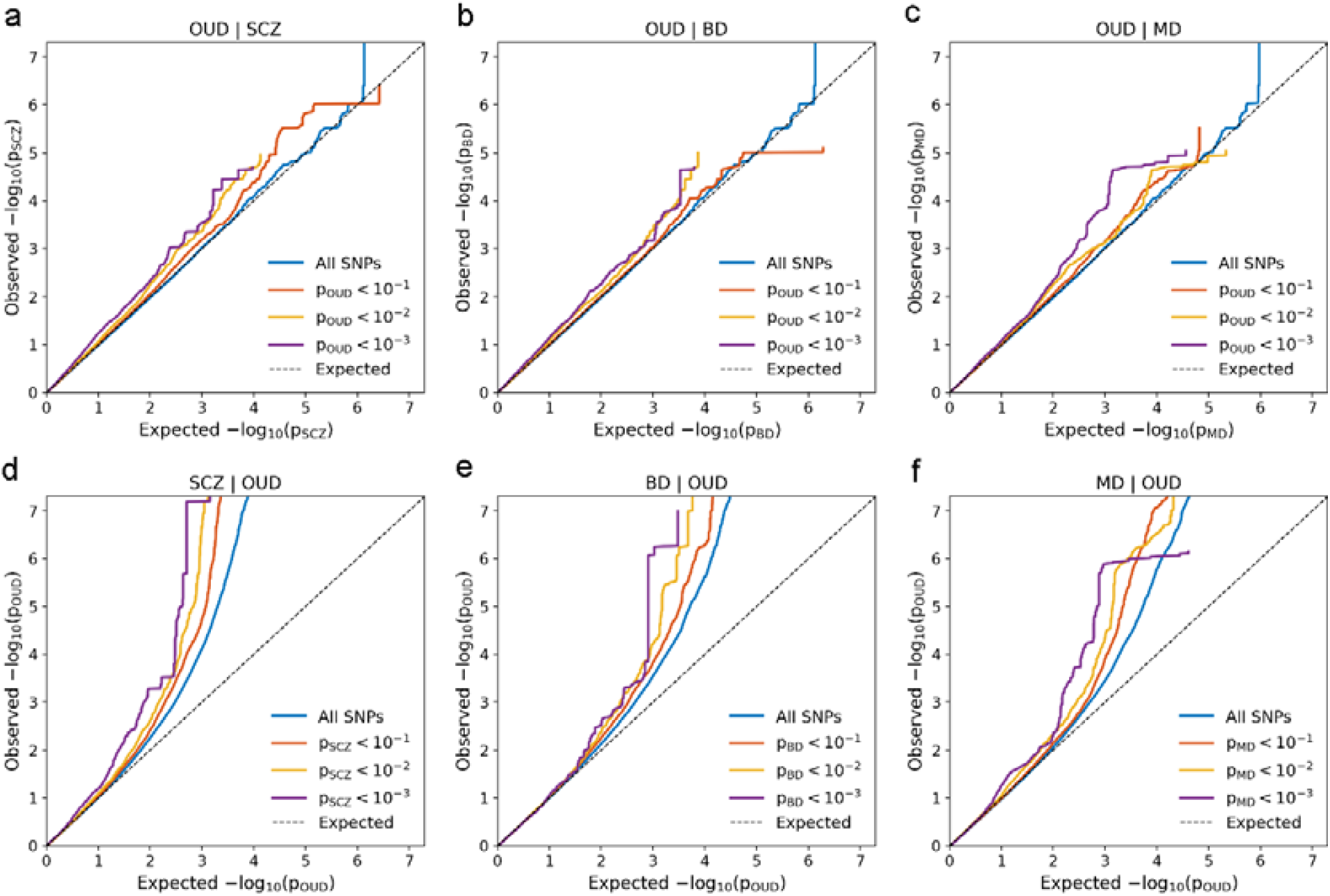
a-c: Conditional Q-Q plots of observed versus expected opioid use disorder (OUD) −log_10_ p-values (corrected for inflation) below the standard GWAS threshold of p < 5×10^−8^ as a function of the significance of the association with schizophrenia (SCZ), bipolar disorder (BD) and major depression (MD), at the level of p ≤ 0.1, p ≤ 0.01, p ≤ 0.001, respectively. d-f: Conditional Q-Q plots of observed versus expected SCZ, BD & MD −log_10_ p-values (corrected for inflation) below the standard GWAS threshold of p < 5×10^−8^ as a function of significance of association with OUD, at the level of p ≤ 0.1, p ≤ 0.01, p ≤ 0.001, respectively. The blue lines illustrate the standard enrichment for all SNPs irrespective of their association p-value in the second phenotype. The dashed line shows the null hypothesis. Successive leftward deflection for declining nominal p-values for a given disorder from the dashed line of no association, indicate that the proportion of non-null SNPs increase with higher levels of association with the other disorders.

### 3.2 CondFDR OUD loci

We applied condFDR to boost discovery of OUD loci conditional on mental disorders, and identified a total of 20 loci for OUD, including 14 novel (condFDR<0.05) (Table 2). This includes 9 loci for OUD conditioned on SCZ (6 novel), 8 loci conditioned on BD (5 novel) and 13 loci conditioned on MD (8 novel), including the locus involving the SNP rs1799971 (*OPRM1*) which is reported in the original OUD GWAS ^21^. The condFDR results are presented in Supplementary Tables 1-3.

**Table 2.** The most strongly associated single nucleotide polymorphisms (SNPs) in genomic loci associated with opioid use disorder (OUD) at conditional false discovery rate (condFDR) < 0.05 given association with schizophrenia (SCZ), bipolar disorder (BD) or major depression (MD) after merging regions < 250 kb apart into a single locus are shown. The table presents chromosomal position (Chr.), conditional false discovery rate (FDR) value, conditioned trait and OUD novelty status. For more details including genes mapped to these loci and the full list of all loci associated with OUD at condFDR < 0.05, see Supplemental Tables 1-3.

| Chr. | Lead SNP | Lead SNP bp | FDR value | Cond. trait | Novelty status, OUD |
| --- | --- | --- | --- | --- | --- |
| 1 | rs67237321 | 55686228 | 3.54E-03 | SCZ, BD, MD | Novel |
| 1 | rs75555622 | 189000000 | 4.24E-02 | BD | Novel |
| 1 | rs78058152 | 237000000 | 1.70E-02 | SCZ, MD | Novel |
| 5 | rs28524171 | 61519672 | 2.05E-02 | MD | Novel |
| 5 | rs10067519 | 166000000 | 3.62E-02 | MD | Novel |
| 6 | rs3777755 | 12159699 | 2.66E-02 | BD | Novel |
| 6 | rs6929812 | 27384520 | 2.66E-02 | MD | MHC |
| 6 | rs442439 | 28573006 | 2.81E-02 | SCZ | MHC |
| 6 | rs1799971 | 154000000 | 4.07E-03 | SCZ, BD, MD | Zhou 2020 <sup>21</sup> |
| 8 | rs35604666 | 93037953 | 3.76E-02 | BD | Novel |
| 9 | rs10761184 | 95698931 | 2.20E-02 | SCZ | Novel |
| 9 | rs864882 | 128000000 | 2.67E-03 | MD | Kember 2021 <sup>39</sup> |
| 10 | rs7088136 | 29159490 | 4.33E-02 | MD | Novel |
| 10 | rs17113901 | 103000000 | 4.04E-02 | SCZ | Kember 2021 <sup>39</sup> |
| 11 | rs1800498 | 113000000 | 4.48E-02 | BD, MD | Kember 2021 <sup>39</sup>, Deak 2022 <sup>43</sup> |
| 11 | rs9919557 | 113000000 | 4.47E-02 | BD, MD | Kember 2021 <sup>39</sup>, Deak 2022 <sup>43</sup> |
| 13 | rs116994084 | 25828354 | 4.41E-02 | MD | Novel |
| 13 | rs2181967 | 31832054 | 3.90E-02 | MD | Novel |
| 14 | rs10145081 | 80243323 | 8.65E-03 | SCZ, MD | Novel |
| 15 | rs62011989 | 54566664 | 1.92E-02 | SCZ | Novel |
| 15 | rs4702 | 91426560 | 9.45E-03 | SCZ, BD, MD | Kember 2021 <sup>39</sup> |
| 19 | rs117623407 | 32204489 | 1.77E-02 | BD | Novel |

### 3.3 Shared loci between OUD and mental disorders (conjFDR)

To identify shared loci between OUD and the psychiatric disorders we performed conjFDR analyses with a threshold at <0.05 (Figure 2). We identified in total seven loci, two of which are novel for OUD and shared with MD at *RN7SKP157* and *B3GALTL* (Table 3). One locus was novel for BD at *DRD2* and one was novel for MD at *PPPC6* (Table 3). For all lead SNPs in the shared loci, the effect directions in OUD and each psychiatric disorder were concordant, i.e., risk of OUD was linked to higher risk of mental illness (Supplementary Tables 4-6). As expected, we found a locus shared between OUD, SCZ and MD located to the MHC region which has previously been implicated in these disorders at the genome wide significant level ^39-41^. This overlapping genetic signal suggests involvement of the immune system in the shared genetic risk of these disorders. However, the extended MHC region has highly complex LD structure spanning a large number of genes. We cannot reliably infer whether this overlapping signal reflects separate or shared loci, or causal genes.

**Figure 2.**
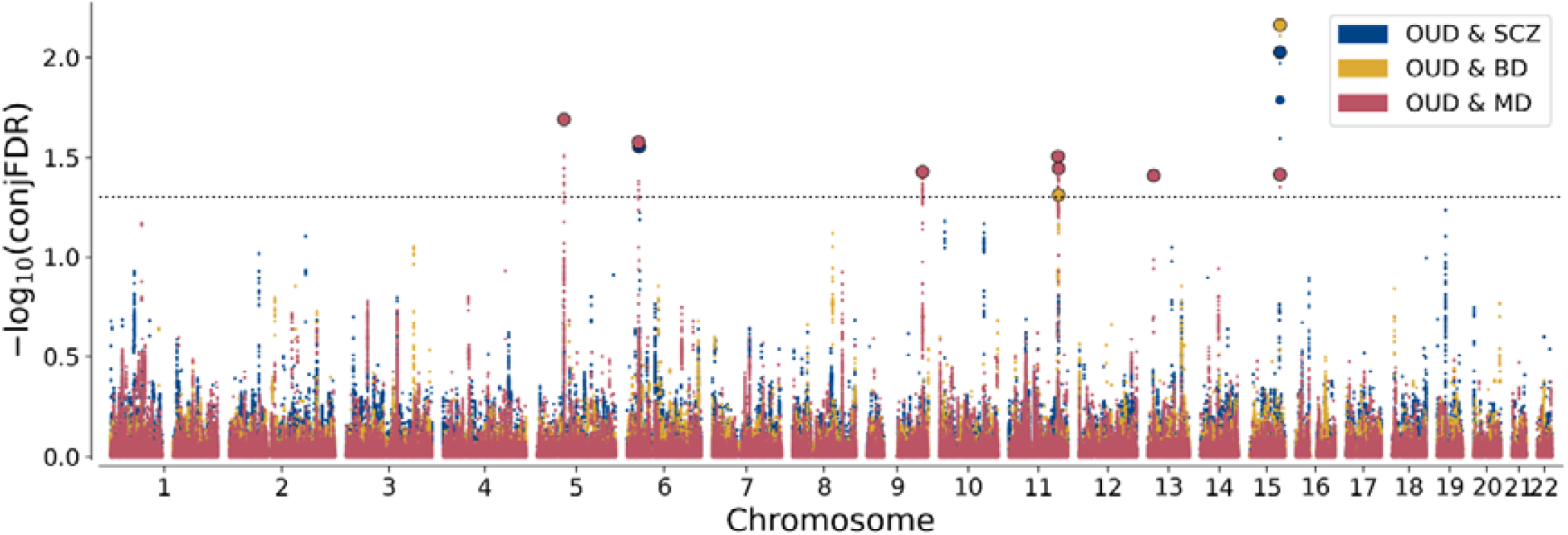
ConjFDR Manhattan plot. Common genetic variants jointly associated between opioid use disorder (OUD) and schizophrenia (SCZ), bipolar disorder (BD) and major depression (MD) at conjunctional false discovery rate (conjFDR) < 0.05. Showing the –log_10_ transformed conjFDR values for each single nucleotide polymorphism (SNP) on the y-axis and chromosomal positions along the x-axis. The dotted horizontal line represents the threshold for significant shared associations (conjFDR=0.05 i.e. −log10[conjFDR]□>1.3). Independent lead SNPs are encircled in black.

**Table 3.**
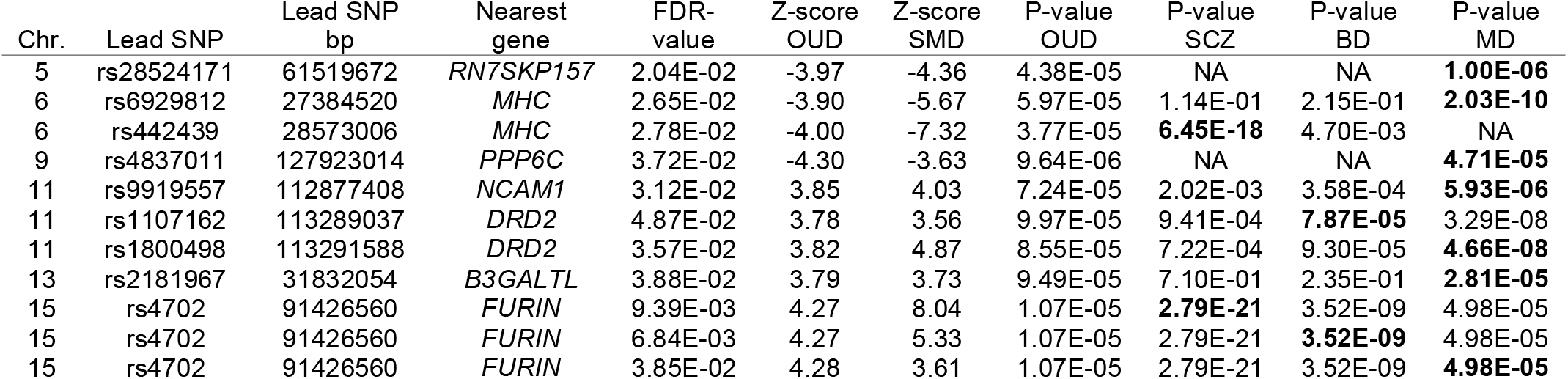
Summary data of shared loci identified between opioid use disorder (OUD) and severe mental disorders (SMDs); schizophrenia (SCZ), bipolar disorder (BD) and major depression (MD) at conjunctional false discovery rate (conjFDR) < 0.05. Bp = base pair; SNP = single nucleotide polymorphism; Chr = chromosome, Bold p-value refers to the listed SMD z-score.

### 3.4 Functional characterization

Functional analyses of candidate SNPs in the loci conditioned on mental disorders with condFDR revealed that most were intergenic and intronic (Supplementary Tables 1-3). After testing for enrichment (e>1) according to functional category exonic (e=2.27, Fisher’s p=0.03) and intronic variants (e=1.15, Fisher’s p=0.03) were significantly enriched for OUD|SCZ. For OUD|BD intergenic (e=1.17, Fisher’s p=5.02E-07) and intronic (e=1.08, Fisher’s p=0.05) variants were significantly enriched. For OUD|MD intronic (e=1.50, Fisher’s p=2.42E-25) variant were significantly enriched. Functional analyses of candidate SNPs in the shared loci identified by conjFDR revealed that most were intergenic and intronic (Supplementary Tables 7-9). After testing for enrichment according to functional category UTR3 (e=5.43, Fisher’s p=0.01) and exonic variants (e=4.99, Fisher’s p=0.01) were significantly enriched for OUD/SCZ. For OUD/BD exonic (e=8.21, Fisher’s p= 9.66E-05) and intronic (e=2.21, Fisher’s p=2.92E-14) variants were significantly enriched. For OUD/MD intronic (e=1.77, Fisher’s p= 3.38E-35) variant were significantly enriched. Mapping of shared loci to genes ^35^ resulted in 98 identified genes shared between OUD and SCZ in two loci (Supplementary Table 10). Thirty of these were left after filtering on two out of three mapping criteria (positional, eQTL, and chromatin interaction information). We identified 19 genes shared between OUD and BD in two loci (Supplementary Table 11). Five of these passed filtering with two out of three mapping strategies. Finally, we identified 145 genes shared between OUD and MD in seven loci (Supplementary Table 12). Thirty-six genes passed the filtering strategy.

### 3.5 Validation analyses

We used the sign concordance test ^34^ to validate the shared loci which produced one of two concordant variants (50%; binomial p=0.75) for SCZ, two of two (100%; binomial p=0.25) concordant variants for BD, six of seven (86%; binomial p=0.06) concordant variants for MD, and four of eight (50%; binomial p=0.64) concordant variants for OUD. We further assessed validity of the lead SNPs in loci identified at conjFDR<0.05 by testing for presence in independent datasets at p<0.05. One lead variant (rs4702 at *FURIN*) was present in the SCZ GWAS of East Asian ancestry ^32^. No lead variants were present in the BD GWAS of European ancestry (https://r5.finngen.fi/). For MD, three of seven lead SNPs (rs28524171 at *RN7SKP157*, rs6929812 within MHC, and rs1800498 at *DRD2*) was validated in the independent MD GWAS of European ancestry ^33^. Finally, no lead variants in the shared loci for OUD and psychiatric disorders, had a p-value <0.05 in the independent AA GWAS dataset on OUD ^21^. A total of four of the OUD risk loci identified by conjFDR were identified in recent larger GWAS on OUD, specifically the *PPP6C* ^42^, *NCAM1* ^39^, *DRD2* ^39^ and *FURIN* ^39,42,43^ loci, supporting the validity of the results.

## 4 DISCUSSION

In the current study, we used the cond/conjFDR analytical method and leveraged cross-trait enrichment between OUD, and SCZ, BD and MD to improve the statistical power for discovery of shared genetic loci to shed light on their genetic relationships. We identified polygenic overlap beyond genetic correlation. Specifically, we discovered 14 novel OUD loci and 7 novel loci jointly associated with OUD and psychiatric disorders (Figure 2, Table 2 and 3). There was a concordant allelic direction of effects in all the shared loci, indicating that these genetic variants increase the risk of both OUD and the respective psychiatric disorders. Several lines of evidence suggest a pleiotropic nature of mental traits and disorders ^44^. Our findings are in line with the observation that multiple genetic variants with small effect sizes influence many traits to different degrees ^17^. The currently identified shared loci explain only a small fraction of the genetic architecture of OUD and severe mental disorders and, thus, the liability to these disorders. We identified 14 novel loci for OUD, but there are still numerous undetected common variants, which will be identified with larger GWAS samples ^17^. In addition, environmental factors, copy number and rare genetic variants play an important role in the aetiology of OUD ^45^, SCZ ^46^, BD ^47^ and MD ^48^. The present results increase our understanding of the underlying genetic variants influencing OUD and the comorbidity between OUD and the severe psychiatric disorders SCZ, BD and MD. These common genetic variants may help generate novel hypotheses about the underlying molecular relationships and their role in the development of psychopathology, whether overlapping loci represent components of discrete clusters of risk (e.g., for substance use disorders) or a general vulnerability to psychiatric symptoms in general ^49^, is still an open question.

We identified a pleiotropic locus on chromosome 15 (lead SNP rs4702) which was shared between OUD, SCZ, BD and MD at conjFDR<0.05. It has previously been associated with SCZ ^40^, BD ^27^ and MD ^41^. This locus was not identified in the original OUD GWAS ^21^, but reached genome-wide significance in subsequent GWAS on OUD, validating this finding ^39,42,43^. The lead SNP rs4702 is shown to influence the expression of its nearest gene *FURIN* in neurons derived from human induced pluripotent stem cells, and influence synaptic function in synergy with other SCZ risk variants ^50^. *FURIN* encodes a cleaving and activating enzyme involved in cleavage of the endogenous opioid proenkephalin ^51^, which is found in most parts of the brain and nervous system ^52^. We discovered a novel OUD locus at chromosome 5 shared with MD, which has previously been associated with both SCZ ^32^ and MD ^53^. The nearest gene to the lead SNP is pseudogene *RN7SKP157*, while the most likely causal genes according to the V2G algorithm were *KIF2A* and *DIMT1*. We also discovered a novel OUD locus at chromosome 13, implicating three genes (*HSPH1, RXFP2* and *B3GALTL*). This locus is shared with MD and has previously been implicated in SCZ ^32^ and MD ^28^. FUMA and V2G both identified the nearest gene and most likely implicated gene as *B3GALTL*, respectively. We identified another OUD risk locus on chromosome 11 which was shared with both BD and MD. The locus is novel for BD and has previously been implicated in OUD ^43^, MD ^41^, as well as for alcohol use disorder ^54^ and SCZ ^40^.The nearest gene to the lead SNP is *DRD2*, while the most likely causal genes according to the V2G algorithm was *ANKK1* and *TTC12*. The gene cluster *TTC12-ANKK1-DRD2-NCAM1* (NTAD) have previously been widely linked to nicotine ^55^, alcohol ^56^ and other drug dependencies ^57^. *NCAM1* has previously been found to be associated with SCZ, BD, MD, alcohol use disorder and cannabis use disorder ^54^. Sequence variants in the *DRD2* gene has been associated with SCZ ^58^, Parkinson’s disease ^59^ and opioid addiction ^60^.

The identified genetic variants seem to support neurobiological hypotheses about the reward system in substance use disorders (SUD) and severe mental disorders (SMDs). The identified loci include variants that may result in changed function of the mesocorticolimbic circuit, influenced by altered metabolism of opioids (*FURIN* variants) and altered dopamine receptor function (*DRD2* variants). Such functional modification of the reward system may lead to an increased risk of SUDs and SMDs, for example via adapted dopamine transmission leading to reduced reward processing ^14^ which may underlie negative psychotic symptoms and depressive mood, and increased susceptibility for exogenous opioids due to elevated dopamine release ^61^ resulting in higher risk of opioid use disorder. Our findings warrant experimental validation to elucidate the pathophysiological mechanisms of the reward circuitry underlying comorbid SUDs and SMDs.

The current study has some limitations. We cannot exclude sporadic, non-systematic sample overlap; however, we calculated linkage disequilibrium score regression genetic covariance intercepts and neither is significant (all p-values>0.05), suggesting no significant effect of overlap between the samples. We included datasets from a diverse set of populations, but larger datasets for other cohorts than Europeans are needed to uncover more of the genetic underpinnings of OUD and SMDs.

To conclude, our study suggests overlapping genetic architecture between OUD and SCZ, BD, and MD, indicating similarities in the genetic architectures of these debilitating disorders. We highlight loci previously associated with SUDs and suggest underlying molecular pathways for OUD. Uncovering the underlying genetics of these disorders can lead to improvements in patient stratification and identification of novel targets for drug development.

## Data Availability

All data produced in the present study are available upon reasonable request to the authors

## ACKNOWLEDGMENTS

We thank the Psychiatric Genomics Consortium, the Million Veteran Program, FinnGen biobank and 23andMe for access to data, and the many people who provided DNA samples. The work was supported by the Research Council of Norway (324499, 324252, 273291, 223273), South-Eastern Norway Regional Health Authority (2019-108), European Union’s Horizon 2020 Research and Innovation Programme grant # 847776 (CoMorMent), and KG Jebsen Stiftelsen (SKGJ□MED□ 021). A.M.D. was supported by NIH grant U24DA041123. This work was performed on the TSD (Tjeneste for Sensitive Data) facilities, owned by the University of Oslo, operated and developed by the TSD service group at the University of Oslo, IT-Department (USIT).

## AUTHOR CONTRIBUTIONS

BH, AAS, OBS, and OAA were responsible for the study concept and design. AAS, KSOC and OAA contributed to the acquisition of summary data. BH, OBS, OAA, and AAS assisted with data analysis and interpretation of findings. AAS prepared figures. BH drafted the manuscript. OBS and OAA provided critical revision of the manuscript for important intellectual content. All authors critically reviewed content and approved final version for publication.

