## Supplemental information for "Genome-wide analyses reveal novel opioid use disorder loci and genetic overlap with schizophrenia, bipolar disorder, and major depression"

**SUPPLEMENTARY METHODS**

**Genome-wide association study data**

*Discovery samples*

Genome-wide association study (GWAS) summary statistics on opioid use disorder (OUD) was obtained from the Million Veteran Program (MVP) [1], a biobank and observational cohort study in the Department of Veterans Affairs (VA) health care system [2]. The MVP cohort enrolment of participants was completed between January 10, 2011, and May 21, 2018, with electronic health record data for OUD diagnosis from October 1, 1999, to February 7, 2018, and blood samples for genetic analyses. Two phases of genotyping were included in the cohort, phase one with 353 948 individuals, 209 020 of whom were non-related European American (EA) individuals. Phase two included 108 416 individuals of whom 67 268 were unrelated EA individuals. Cases were defined as either one inpatient ICD-9 or ICD-10 code for OUD or two outpatient codes. In phase one there were 6367 EA individuals and in phase two 2162 EA individuals with OUD. Opioid-exposed controls were selected by taking all MVP participants and excluding participants with exposure of an opioid prescription for < seven consecutive days and VA follow up < six months after baseline, participants with cancer diagnosis before or after baseline, with opioid daily dosage at baseline of more than 90 mg morphine equivalents, OUD treatment or diagnosis at baseline. After this filtering a latent growth mixture model was used to classify opioid dose trajectories, measured by morphine equivalent daily dose. Individuals were assigned to one of four groups (low, moderate, escalating and rapidly escalating) of most likely belonging. The low-dose trajectory group without OUD diagnosis at follow-up were designated controls, yielding 55 429 EA controls in phase one and 15 771 EA controls in phase two. An Affymetrix Biobank Array was used for genotyping in the MVP cohort. [3] has previously described imputation and quality control for the MVP phase one. In short, EAGLE [4] was used to perform pre-phase chromosome imputation, and to impute genotypes with 1000 Genomes Project phase three data as reference panel [5], Minimac [6] was used. When demographic information was missing and when genotypic and phenotypic sex did not match, they were removed. One subject from related pairs were also randomly removed (kinship coefficient threshold = 0.0884). To deal with network-like relationships between three or more individuals a greedy algorithm was implemented, leaving 353 948 individuals, 209 020 of whom were non-related EA individuals, for subsequent analyses. Phase two dataset quality control follow a similar process as phase one and yielded 107 438 subjects for subsequent analyses. Next, principal component analysis (PCA) was run on 74 827 common SNPs with a mean allele frequency (MAF) of > 0.05 shared by MVP and the 1000 Genomes phase three reference panels by FastPCA [7]. The first ten principal components (PC)s were used for clustering of participants into five reference populations (African, admixed American, East Asian, European, and South Asian) by their Euclidean distance from the closet cluster. Outliers were removed in the European and African clusters after running another PC analysis. PCs > three standard deviations from the mean were removed, leaving 67,268 EA unrelated subjects. Imputation in the MVP phase two was similar to phase one. A posterior probability of ≥0.9 was used as a threshold for imputed genotypes and were transferred to best-guess genotypes. The rest were treated as missing genotype calls. 5.1 million SNPs in EA and 6.9 million in AA clusters with INFO scores ≥0.7, genotype call rates or best-guess rates >0.95, Hardy-Weinberg Equilibrium p-value >1×10−6, and minor allele frequency (MAF) >0.01 were retained for analysis. GWAS was then run with PLINK [8] using logistic regression, correcting for age, sex, and the first ten principal components. OUD case-control GWAS results from Yale-Penn (data collected between February 14, 1999, and April 1, 2017) and SAGE (data collected from 1990 to 2007) were also used. All subjects were opioid exposed. For EAs, Yale-Penn one contains 1,043 cases and 294 controls. Yale-Penn two contains 724 cases and 243 controls. Yale-Penn three contains 54 cases and 44 controls. The Study of Addiction: Genetics and Environment (SAGE) [9] contains 194 cases and 382 controls. All together there were 10,544 EA cases and 72,163 EA opioid-exposed controls. GEMMA8 [10] were used for association analyses. Relatedness and correction for age, sex, and the first ten PCs were taken into account. Please refer to the original publication for further details [1].

The schizophrenia (SCZ) dataset was obtained from the Psychiatric Genomics Consortium (PGC) Schizophrenia Work Group and includes 90 core case/control and family-based cohorts with individual-level genotype data processed in a uniform pipeline [11]. Phenotypic data was quality controlled by a systematic review of data collection methods and procedures at each site, including only studies fulfilling these criteria. Cases and controls were matched by geography and ancestry, and harmonizing quality control of the cohorts were done separately in accordance with PGC standards [12]. We excluded all participants of East Asian, African American, and Latino ancestry to avoid confounding due to differences in inheritance patterns across ancestries. Our main analysis comprised 53,386 European cases of SCZ and schizoaffective disorder and 77,258 European controls. Diagnostic strategy included consensus diagnosis, research diagnostic interview, review of medical records or a mixed strategy. Exclusion details can be found in the original publication [11]. Quality control of the cohorts were done separately, in brief SNP missingness > 0.05 (before sample removal); subject missingness > 0.02; autosomal heterozygosity deviation (|Fhet| > 0.2); SNP missingness > 0.02 (after sample removal); difference in SNP missingness between cases and controls > 0.02; and SNP Hardy–Weinberg equilibrium (HWE: P < 10−6 in controls or P < 10−10 in cases) were filtered out. For the family-based cohorts individuals with >10,000 Mendelian errors and SNPs with >4 Mendelian errors were excluded. PCA was performed for cohorts separately using SNPs with high imputation quality (INFO > 0.8), low missingness (<1%), MAF > 0.05 and in relative linkage equilibrium (LD) after two iterations of LD pruning (r2 < 0.2, 200 SNP windows). Areas with known long-range LD (MHC and chr8 inversion) were removed. EAGLE 2 [4] or MINIMAC3 [13] was used for genotype prephasing/imputation in case control cohorts. The Haplotype Reference Consortium (HRC) reference panel was used for imputation [14]. Family-based trios were phased with SHAPEIT 3 [15]. SNPs present in all cohorts were used for relatedness testing with PLINK v.1.945 where pairs of subjects with $\hat{\pi}$> 0.2 were identified and one member of each pair removed at random, prioritizing retention of cases and trio members over case–control members. Using PLINK, an additive logistic regression model was leveraged as basis for association testing. The first four PCAs were included as a default and then every PCA that was nominally significant to case control status. In trios PCA was only used to remove extreme ancestry outliers. Finally, the results were meta-analysed by a standard inverse-weighted fixed effects model. Please see original publication for further details [11].

The bipolar disorder (BD) dataset was obtained from the PGC Bipolar Disorder Working Group [16]. The study comprises 57 cohorts from Europe, North America, and Australia, includes 41,917 cases and 371,549 controls all of European ancestry. Cases were diagnosed via the Diagnostic and Statistical Manual of Mental Disorders (DSM)-IV, International Classification of Diseases (ICD)-9 or -10 and had to qualify for a lifetime diagnosis of BD, set by structured diagnostic instruments administered by trained interviewers, clinician-administered checklists, or medical record review. In most cohorts controls were randomly selected from the population and screened for lifetime psychiatric disorders. Quality control for retention of SNPs and subjects included SNP missingness < 0.05 (before sample removal), SNP missingness < 0.02 (after sample removal), subject missingness < 0.02, and difference in SNP missingness between cases and controls < 0.02, and a SNP Hardy–Weinberg equilibrium test (P > 10 × 10−10 in psychiatric cases and P > 10 × 10−6 in controls). One of each pair of related individuals ($\hat{\pi}$ > 0.2) was excluded. Imputation of genotypes was performed by a stepwise approach using Eagle v 2.3.5 [4] and Minimac3 [13] to the HRC reference panel v1.0 [14]. An additive logistic regression model in PLINK v1.90 [8] was used to run the GWAS, covarying for the five first PCs and more where needed. See original publication for further details [16].

The dataset for major depression (MD) was obtained from the Psychiatric Genetics Consortium (PGC) [17]. 331,374 individuals were left after excluding 79,990 individuals that were outliers based on heterozygosity, had a variant call rate <98%, or were not recorded as “white British”. 131,790 individuals were excluded based on a shared relatedness of up to the third-degree using kinship coefficients (>0.044) calculated using the KING toolset [18]. One member was subsequently added back in each group of related individuals by creating a genomic relationship matrix and selected individuals with a genetic relatedness less than 0.025 with any other participant (n = 55,745). Variants with a call rate < 98%, a minor allele frequency <0.01, deviation from Hardy–Weinberg equilibrium (P < 10−6) or an imputation accuracy (INFO) score < 0.1 were removed. Broad depression (113,769 cases, 208,811 controls), Probable Major depressive disorder (MDD) (30,603 cases, 143,916 controls) and ICD-coded MDD (8,276 cases, 209,308 controls) were included in the dataset. Participants with bipolar disorder, schizophrenia or personality disorder using self-declared data or ICD codes from hospital admission records; and participants who reported having a prescription for an antipsychotic medication during a verbal interview were excluded. Further exclusions were applied to all control individuals if they had a diagnosis of a depressive mood disorder from hospital admission records, had reported having a prescription for antidepressants or had self-reported depression. Genotype date was imputed with IMPUTE4 using the HRC reference panel [14]. BGENIE [19] was used for linear association testing of the effect of each variant. Covariates included sex, age, genotyping array and eight PCs. Odd’s ratios of variants associated with depression were obtained by logistic regression conducted in Plink 1.90b4 [8] using the same covariates as the linear model. Further details are available at the original publication [17].

*Validation samples*

We used the AA cohort of the MVP datasets for validation analyses for OUD [1]. After quality control and imputation, as described above for the EA cohorts, phase one of the MVP contains 57 340 and phase two 18 214 unrelated AA individuals. Cases and controls were defined as for the EA cohorts. Phase one contained 3151 AA individuals and phase two 881 AA individuals with OUD. There were 20 254 AA controls in phase one and 5775 AA controls in phase two. For the Yale-Penn and SAGE cohort for AAs, Yale-Penn one contains 831 cases and 573 controls. Yale-Penn two contains 349 cases and 274 controls. For AA individuals only the two first phases of the Yale-Penn and SAGE cohorts were included due to the low number of AA individuals in phase three (Yale-Penn three has seven cases and SAGE has 105 cases and 158 exposed controls). To assess validation in OUD 5,212 cases and 26,876 controls of AA ancestry were used in total from the MVP and Yale-Penn and SAGE cohorts [1]. See EA cohort and original publication for description of diagnostic, imputation and association testing [1].

The SCZ GWAS dataset was obtained from the PGC [20]. It contains 22,778 cases and 35,362 controls of East Asian ancestry [20]. Japanese, Korean, Indonesian, and Han Chinese populations were included. The samples were based on 18 case-control studies (21 709 cases 33 633 controls) and two family-based association studies (1699 parent affected-offspring trios). DSM-IV and Diagnostic interview for genetic studies (DIGS) were used for diagnostics. Phenotypic data quality control was verified by the same procedures as in the SCZ GWAS used as discovery dataset, also conducted by the PGC [11]. Genotypes were prephased and imputed with SHAPEIT [21] and IMPUTE2 [22] using the 1000 Genomes Project reference panel [23]. Additional processing for parent affected-offspring trios was carried out such that case/controls were identified and imputed. Quality control procedures were carried out as part of the RICOPILI pipeline [24]. PCA was conducted across samples via imputed best-guess genotypes to identify and remove overlapping samples across datasets, cryptic related samples, and population outliers. Eight principal components that were associated with case-control status were included in univariate logistic regression as covariates to control for the population stratification in each sample. Q-Q plots showed that the population structure was well controlled. SNPs missing in ≥ 5 % of subjects were excluded. Association analysis was carried out for each sample using PLINK [25] and genotype dosage from imputation. Only variants with imputation INFO ≥ 0.6 and a MAF ≥ 1% were included in the analysis. Fixed-effect meta-analysis [26], weighted by inverse variance, was then used to combine association results across samples. For further details, please see the original publication [27].

The BD GWAS summary statistics were obtained from the FinnGen Data Freeze 5 ([https://r5.finngen.fi/](https://emea01.safelinks.protection.outlook.com/?url=https%3A%2F%2Fr5.finngen.fi%2F&data=04%7C01%7C%7Cc1a2eee026a74e63f27608d92506b887%7C84df9e7fe9f640afb435aaaaaaaaaaaa%7C1%7C0%7C637581533062698830%7CUnknown%7CTWFpbGZsb3d8eyJWIjoiMC4wLjAwMDAiLCJQIjoiV2luMzIiLCJBTiI6Ik1haWwiLCJXVCI6Mn0%3D%7C1000&sdata=7Z09JT%2BfJn2HFuHXvQvaOEpXCiBbE2%2FhxFvkZW%2BjlH0%3D&reserved=0)). The dataset comprises 4,501 cases and 192,220 controls. BD diagnostics were made with the ICD-8, -9 and -10. Genotyping was done on Illumina and Affymetrix arrays (Illumina Inc., San Diego, and Thermo Fisher Scientific, Santa Clara, CA, USA), and genotype calls were made with GenCall or zCall for Illumina and AxiomGT1 for Affymetrix data. Individuals with ambiguous gender, high genotype missingness (>5%), excess heterozygosity (+-4SD) and non-Finnish ancestry were excluded, as well as all variants with high missingness (>2%), low Hardy–Weinberg equilibrium *p*-value (<1e-6) and minor allele count (MAC < 3). GWAS was run with SAIGE, a mixed model logistic regression R/C++ package, version 0.36.3.2 (<https://github.com/weizhouUMICH/SAIGE/tree/finngen_r5_jk>). Covariates included sex, age, ten principal components and genotyping batch.

The independent summary statistics for MD was obtained from the consumer genetics company 23andMe [28]. Data included in this cohort was collected in January 2015 and January 2016 [29, 30] and included self-reported 75,607 cases (clinical diagnosis or treatment of depression) of European ancestry and 231,747 controls (no history of depression) of European ancestry, around 58% male and an average age of 46 (approx. 95% of the group between the ages of 20 and 80). Most participants are from the United States, with the next largest groups from Canada and Europe. Genotyping was performed with variants of the Illumina HumanHap550+ BeadChip, Illumina OmniExpress+ BeadChip or a fully custom array. Samples that failed to reach a call rate of 98.5% were reanalysed. Imputation and prephasing was done with SHAPEIT2 [31] and a tool developed internally at 23andMe, Finch [32] against the 1000 Genomes Project reference panel phase one. Prior to imputation phased chromosomes were split into segments of no more than 10,000 genotyped SNPs, with overlaps of 200 SNPs. SNPs with Hardy–Weinberg equilibrium P value <1 × 10−20, call rate <95%, or large allele frequency discrepancies in comparison to European 1000 Genomes Project reference data were excluded. Each phased segment was imputed against all-ancestry 1000 Genomes Project haplotypes (excluding monomorphic and singleton sites) using Minimac2 [33]. The analysis was restricted to individuals who had >97% European ancestry, as determined through an analysis of local ancestry [34]. PCA was used to characterize residual population structure. Principal components were computed using 82,654 SNPs that were genotyped on all 23andMe array designs, with Hardy–Weinberg P value >1 × 10−40, MAF >0.01, and call rate >99%, excluding regions of extended long-range LD. In the GWAS association test results was computed by logistic regression assuming additive allelic effects. For tests using imputed data, the imputed dosages rather than best-guess genotypes were used. Covariates included age, sex, and the top five principal components to account for residual population structure. For quality control of genotyped GWAS results, SNPs that were only genotyped on the two first genotype platforms were removed due to small sample sizes. SNPs on the mitochondrial or Y chromosome, was also removed. Using family trio data, SNPs that failed a test for parent–offspring transmission was marked; specifically, regression of the child's allele count against the mean parental allele count was marked and SNPs with fitted β<0.6 and P <1 × 10−20 for a test of β<1 was done. We removed SNPs with a Hardy–Weinberg P value <1 × 10−20 in Europeans or a call rate of <90%. SNPs with average r2 <0.5 or minimum r2 <0.3 in any imputation batch were removed, as well as SNPs that had strong evidence of an imputation batch effect. For further detail, please see the original publication [28].

**Conditional Q-Q plots and cross-trait enrichment**

Under large-scale testing paradigms, such as GWAS, quantitative estimates of likely true associations can be obtained from the distributions of summary statistics [35, 36]. One common method for visualizing the enrichment of statistical association relative to that expected under the global null hypothesis is through Q-Q plots of nominal p-values obtained from GWAS summary statistics. The Q-Q curve has as the y-ordinate the nominal p-value, denoted by “p”, and as the x-ordinate the corresponding value of the empirical cumulative distribution function (cdf), denoted by “q”. Under the global null hypothesis, the theoretical distribution is uniform on the interval [0,1]. In the presence of all null relationships, nominal p-values form a straight line on a Q-Q plot when plotted against the empirical distribution. Leftward deflections of the observed distribution from the projected null line reflect increased tail probabilities in the distribution of test statistics (z-scores) and consequently an over-abundance of low p-values compared to that expected by chance, also named ‘enrichment’. To emphasize tail probabilities of the theoretical and empirical distributions, the log_10_ p is commonly plotted against the -log_10_ q.

Conditional Q-Q plots are constructed by creating subsets of SNPs based on levels of an auxiliary measure for each SNP, and computing Q-Q plots separately for each level [37]. If SNP enrichment is captured by variation in the auxiliary measure, this is expressed as successive leftward deflections in a conditional Q-Q plot as levels of the auxiliary measure increase. The enrichment can be directly interpreted in terms of the true discovery rate (1−FDR) (see below) [38]. Cross-trait enrichment exists if the proportion of SNPs associated with a phenotype increases as a function of the strength of the association with a secondary phenotype. We constructed conditional Q-Q plots of empirical quantiles of nominal -log_10_ p-values for SNP association for all SNPs, and for subsets (strata) of SNPs determined by the nominal p-values of their association with the conditional phenotypes, and vice versa. Specifically, we computed the empirical cumulative distribution of nominal p-values for a given phenotype for all SNPs and for SNPs with significance levels below the indicated cut-offs for the conditional phenotypes (-log_10_(p) ≥ 1, -log_10_(p) ≥ 2, log_10_(p) ≥ 3 corresponding to p < 0.1, p < 0.01, p < 0.001 respectively). The nominal p-values (– log_10_(p)) are plotted on the y-axis, and the empirical quantiles (–log_10_(q), where q=1-cdf(p)) are plotted on the x-axis. To assess for polygenic effects below the standard GWAS significance threshold, we focused the conditional Q-Q plots on SNPs with nominal –log_10_(p) < 7.3 (corresponding to p > 5x10^-8^).

**Conditional and conjunctional false discovery rate**

The ‘enrichment’ seen in conditional Q-Q plots can be directly interpreted in terms of a Bayesian true discovery rate (1 – FDR) [38]. More specifically, for a given p-value, under a simple two-group (null and non-null) model, Bayes rule gives the posterior probability of being null as

FDR(p) = π_0_F_0_ (p) / F(p),	[1]

where π_0_ is the proportion of null SNPs, F_0_ is the cdf of the null SNPs, and F is the cdf of all SNPs, both null and non-null [35]. Here, we assume the SNP p-values are *a priori* independent and identically distributed. Under the null hypothesis, F_0_ is the cdf of the uniform distribution on the unit interval [0,1], so that Eq. [1] reduces to

FDR(p) = π_0_ p / F(p),	[2]

F can be estimated by the empirical cdf q = N_p_ / Ν, where N_p_ is the number of SNPs with p-values less than or equal to p, and N is the total number of SNPs. Replacing F by q in Eq. [2], we get

Estimated FDR(p) = π_0_ p / q,	[3]

which is biased upwards as an estimate of the FDR.[46] Replacing π_0_ in Equation [3] with unity gives an estimated FDR that is further biased upward;

q* = p/q,	[4]

If π_0_ is close to one, as is likely true for most GWASs, the increase in bias from Eq. [3] is minimal.

The quantity 1 – p/q, is therefore biased downward, and hence a conservative estimate of the TDR. Referring to the Q-Q plots, we see that q* is equivalent to the nominal p-value divided by the empirical quantile, as defined earlier. We can thus read the FDR estimate directly off the Q-Q plot as

-log_10_(q*) = log_10_(q) – log_10_(p),	[5]

i.e., the horizontal shift of the curves in the Q-Q plots from the expected line x = y, with a larger shift corresponding to a smaller FDR. This is illustrated in Figure 1. To estimate the conditional FDR of a given SNP, we repeat the above procedure for a subset of SNPs with p-values in the secondary GWAS equal to or lower than that observed for the given SNP. Formally, this is given by

FDR(p_1_|p_2_) = π_0_ (p_2_)p_1_/ F(p_1_|p_2_),	[6]

where p_1_ is the p-value for the first phenotype, p_2_ is the p-value for the second, F(p_1_ | p_2_) is the conditional cdf, and π_0_ (p_2_) the conditional proportion of null SNPs for the first phenotype given that p-values for the second phenotype are p_2_ or smaller. The conditional FDR framework is closely related to the stratified FDR method developed by [39]. Whereas they propose computing FDR separately conditional on membership in pre-defined discrete strata of p-values, here, we condition the estimated FDR on a continuous random variable, the SNP p-values with respect to a second phenotype.

To identify SNPs jointly associated with two phenotypes using conjunctional FDR, the conditional FDR procedure is repeated after inverting the roles of the primary and secondary phenotypes. Similar to previous conjunction tests for p-value statistics [47], the conjunctional FDR estimate is defined as the maximum of both conditional FDR values, which minimizes the effect of a single phenotype driving the common association signal. Formally, the conjunctional FDR is given by

FDR_Phenotype1&Phenotype2_ (p1, p2) = π0 F0(p1, p2) / F(p1, p2) + π1 F1(p1, p2) / F(p1, p2) + π2 F2(p1, p2) / F(p_1_, p_2_),	[7]

where π_0_ is the *a priori* proportion of SNPs null for both phenotypes simultaneously and F_0_(p_1_, p_2_) is the joint null cdf, π_1_ is the *a priori* proportion of SNPs non-null for the first phenotype and null for the second with F_1_(p_1_, p_2_) the joint cdf of these SNPs, and π_2_ is the *a priori* proportion of SNPs non-null for the second phenotype and null for the first, with joint cdf F_2_(p_1_, p_2_). F(p_1_, p_2_) is the joint overall mixture cdf for all phenotype 1 and 2 SNPs.

Conditional empirical cdfs provide a model-free method to obtain conservative estimates of Eq (7). This can be seen as follows. Estimate the conjunction FDR by

Estimated FDR_Phenotype1&Phenotype2_ =

max {Estimated FDR_Phenotype1|Phenotype2_, Estimated FDR_Phenotype2|Phenotype1_}, [8]

where Estimated FDR_Phenotype1|Phenotype2_ and Estimated FDR_Phenotype2|Phenotype1_ are conservative (upwardly biased) estimates of Eq. [6]. Thus, Eq (8) is a conservative estimate of max {p_1_/F(p_1_| p_2_), p_2_/F(p_2_|p_1_)} = max{p_1_F_2_(p_2_)/F(p_1_, p_2_), p_2_F_1_(p_1_)/F(p_1_, p_2_)}, with F_1_(p_1_) and F_2_(p_2_) the marginal non-null cdfs of SNPs for phenotype 1 and 2, respectively. For enriched samples, p-values will tend to be smaller than predicted from the uniform distribution, so that

F_1_(p_1_) ≥ p_1_ and F_2_(p_2_) ≥ p_2_. Then

max {p_1_F_2_(p_2_) / F(p_1_, p_2_), p_2_F_1_(p_1_) / F(p_1_, p_2_)}

≥ [π_0_ + π_1_ + π_2_] max{p_1_F_2_(p_2_) / F(p_1_, p_2_), p_2_F_1_(p_1_) / F(p_1_, p_2_)}

≥ [π_0_p_1_p_2_ + π_1_p_2_F_1_(p_1_) + π_2_p_1_F_2_(p_2_)] / F(p_1_, p_2_).

Under the assumption that SNPs are independent if one or both are null, reasonable for disjoint samples, this last quantity is precisely the conjunctional FDR given in Eq (7). Thus, Eq (8) is a conservative model-free estimate of the conjunctional FDR.

**Detection of genetic variants using conditional and conjunctional FDR**

The FDR can be interpreted as the probability that a SNP is null given that its p-value is as small as or smaller than its observed p-value. The conditional FDR (condFDR) is an extension of the standard FDR, which incorporates information from GWAS summary statistics of a second phenotype to adjust its significance level. The condFDR is defined as the probability that a SNP is null in the first phenotype given that the p-values in the first and second phenotypes are as small as or smaller than the observed ones. The condFDR estimates are obtained for each nominal SNP p-value in the primary phenotype after computing the stratified empirical cdfs of the nominal p-values [39, 40]. The separate strata are determined by the relative enrichment of SNP associations as a function of increased nominal SNP p-values in a secondary phenotype. The standard FDR framework derives from a model that assumes that the distribution of test statistics in a GWAS can be formulated as a mixture of null and non-null effects, with true associations having more extreme test statistics than false associations on average. Ranking SNPs by the standard FDR or by p-values gives the same ordering of SNPs. In contrast, if the primary and secondary phenotypes are related genetically, the condFDR reorders SNPs and results in a different ranking than that based on p-values alone. The conjunctional FDR (conjFDR) is defined as the posterior probability that a SNP is null for either phenotype or both simultaneously, given that its p-values for association with both phenotypes are as small as or smaller than the observed p-values [41-45]. A conservative estimate of the conjFDR is given by the maximum condFDR for a given SNP after repeating the condFDR procedure for both traits and inverting their roles [37]. If summary statistics for the same SNP were not available in both of the discovery datasets, overlapping SNP associations could not be assessed using conjFDR analysis.

**Functional annotation ang gene prioritization**

Using FUMA [48], an online annotation platform (http://fuma.ctglab.nl/), we functionally annotated all candidate SNPs in the genomic loci with a conjFDR value <0.10 and having an r2
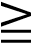
0.6 with one of the independent significant SNPs in the identified loci. SNPs were annotated with Combined Annotation Dependent Depletion (CADD) scores [49], RegulomeDB scores [50], and chromatin states [51, 52]. The CADD score is a deleterious score of variants computed by integrating 63 functional annotations [49]. The higher the score, the more deleterious. A CADD score above 12.37 is the threshold to be potentially pathogenic [49]. The RegulomeDB score is a categorical score to guide interpretation of regulatory variants [50]. It is based on information from expression quantitative trait locus (eQTL) and chromatin marks, ranging from 1a to 7 with lower scores indicating an increased likelihood of having a regulatory function. Scores are as follows: 1a=eQTL + Transcription Factor (TF) binding + matched TF motif + matched DNase Footprint + DNase peak; 1b=eQTL + TF binding + any motif + DNase Footprint + DNase peak; 1c=eQTL + TF binding + matched TF motif + DNase peak; 1d=eQTL + TF binding + any motif + DNase peak; 1e=eQTL + TF binding + matched TF motif; 1f=eQTL + TF binding / DNase peak; 2a=TF binding + matched TF motif + matched DNase Footprint + DNase peak; 2b=TF binding + any motif + DNase Footprint + DNase peak; 2c=TF binding + matched TF motif + DNase peak; 3a=TF binding + any motif + DNase peak; 3b=TF binding + matched TF motif; 4=TF binding + DNase peak; 5=TF binding or DNase peak; 6=other; 7=Not available.[50] The chromatin state represents the accessibility of genomic regions (every 200bp) with 15 categorical states predicted by a hidden Markov model based on 5 chromatin marks for 127 epigenomes in the Roadmap Epigenomics Project [51]. A lower state indicates higher accessibility, with states 1-7 referring to open chromatin states. We annotated the minimum chromatin state across tissues to SNPs. The 15-core chromatin states as suggested by Roadmap are as follows: 1=Active Transcription Start Site (TSS); 2=Flanking Active TSS; 3=Transcription at gene 5’ and 3’; 4=Strong transcription; 5= Weak Transcription; 6=Genic enhancers; 7=Enhancers; 8=Zinc finger genes & repeats; 9=Heterochromatic; 10=Bivalent/PoisedTSS; 11=Flanking Bivalent/Poised TSS/Enh; 12=Bivalent Enhancer; 13=Repressed PolyComb; 14=Weak Repressed PolyComb; 15=Quiescent/Low. Standardized SNP effect sizes were calculated for the most impactful SNPs by transforming the sample size-weighted meta-analysis *Z* score, as described by Zhu et al [52].

Further, we used FUMA [48] to map the candidate SNPs to genes using two of three gene-mapping strategies, specifically their physical position, eQTL functionality and chromatin interactions. Positional mapping links SNPs to genes based on their physical proximity (i.e., within a 10kb window). eQTL mapping matches cis-eQTL SNPs to genes whose expression is associated with allelic variation at the SNP level. We assessed eleven eQTL databases implemented in FUMA which include eQTL information from multiple human tissue types including several brain regions [(http://fuma.ctglab.nl/tutorial#eQTLs)](http://fuma.ctglab.nl/tutorial#eQTLs). The eQTL analyses were corrected for multiple comparisons using an FDR threshold of 0.05. Chromatin interaction mapping links SNPs to genes based on three-dimensional DNA–DNA interactions between each SNP’s genomic region and nearby or distant genes. To increase the likelihood that the prioritized genes from chromatin interaction mapping have a plausible biological function, we only selected interactions where one region overlapped with a predicted enhancer region in any of the 111 tissue/cell types from the Roadmap Epigenomics Project and the other region was located in a gene promoter region (from 250 bp upstream to 500 bp downstream of the transcription start site) as predicted by the Roadmap Epigenomics Project [51]. FUMA contains Hi-C data of more than 21 tissue/cell types including human brain tissue (https://fuma.ctglab.nl/tutorial#chromatin-interactions). An FDR of 1 x 10^-6^ was used to define significant chromatin interactions, as suggested by Schmitt et al (2016) [53].

To complement FUMA we used V2G, a tool developed by Open Targets Genetics (OTG) [54]. V2G prioritizes genes according to the highest overall V2G score. For each genetic variant, the overall V2G score aggregates differentially weighted evidence of variant–gene associations from several data sources, including molecular cis-QTL data (for example, cis-protein QTLs from [55], cis-eQTLs from GTEx v.7 and so on), interaction-based datasets (for example, promoter capture Hi-C), genomic distance and variant effect predictions (VEP) from Ensembl. See (<https://genetics-docs.opentargets.org/our-approach/data-pipeline>) for a detailed description of the evidence sources and weights used [54]. For complementarity and uniformity for gene selection, we investigated each lead variant and identified the positionally nearest gene with FUMA. We then included it in addition to all genes with overall higher V2G scores from Open Targets V2G for further analysis.

**SUPPLEMENTARY FIGURES**

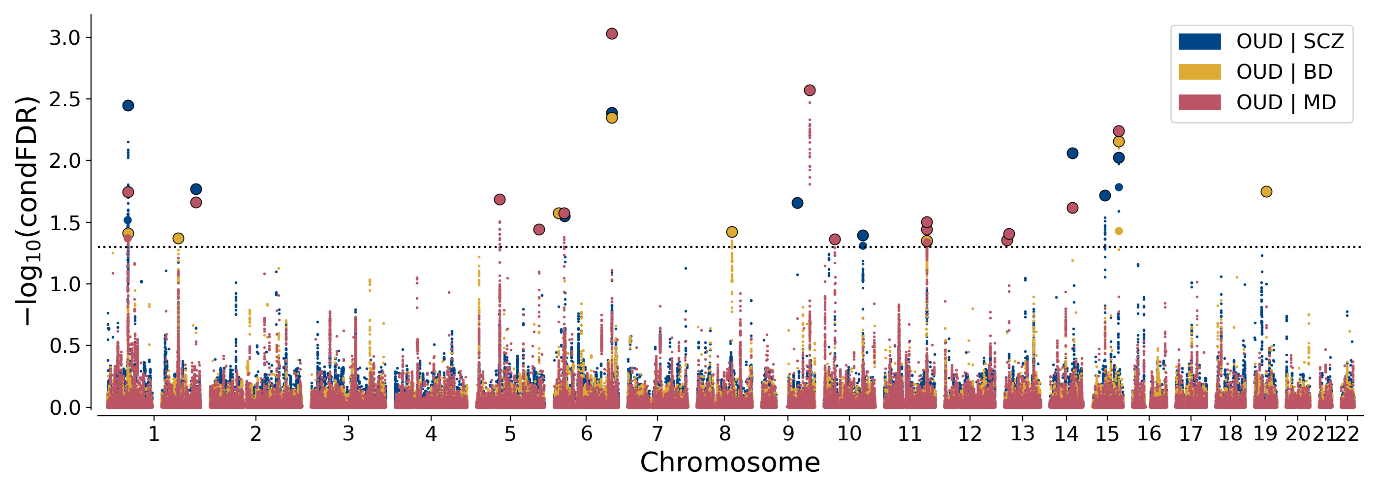
Figure S1. Conditional False Discovery rate (FDR) Manhattan plot for opioid use disorder (OUD) conditioned on schizophrenia (SCZ), bipolar disorder (BD) and major depression (MD). Single-nucleotide polymorphisms (SNPs) with conditional −log10 (FDR) higher than 1.3 (horizontal dotted line) (i.e., conditional FDR < 0.05) are shown with large points. A black line around the large points indicates the most significant SNP in each linkage disequilibrium block. For details, see Supplementary Tables 1-3.
